# What child and adolescent psychiatry in France for the second quarter of the 21^st^ century? An AI-assisted qualitative action research study

**DOI:** 10.1101/2024.07.08.24310059

**Authors:** Bruno Falissard, Pauline Espi, Alexandra Rouquette

**Affiliations:** CESP, INSERM U1018, Université Paris-Saclay, UVSQ, AP-HP, Paris, France; Service de Psychopathologie de l’Enfant et de l’Adolescent, Hospices Civils de Lyon; Pôle de Psychiatrie de l’Enfant et de l’Adolescent, Centre Hospitalier le Vinatier

**Keywords:** Child and Adolescent Psychiatry, Action research, health care system, France

## Abstract

French Child and Adolescent Psychiatry (CAP) faces significant issues, primarily due to an overwhelming increase in demand and insufficient capacity. In response, the French Society for Child and Adolescent Psychiatry and Allied Professions (SFPEADA) initiated an action research project in June 2023 aimed at reimagining the future of CAP in France for the second quarter of the 21st century. Employing a holistic qualitative methodology that merges bottom-up and top-down approaches, the project progressed through four phases: interviews with informed individuals, consultations with trade unions or associations, synthesis of findings using thematic analysis and AI technologies, and public dissemination via a symposium at the ministry of health. The project identified 5 main themes: “CAP and Society”, “Knowledge Integration”, ”Healthcare Delivery”, “Caregivers”, ”System Organization”. This initiative underscores the importance of a collaborative, multidisciplinary approach to address the important needs of child and adolescent mental health in France, advocating for significant systemic changes to enhance CAP’s efficacy and accessibility.

## Introduction

It is now generally accepted that there is a global mental health crisis (1) particularly among children and adolescents (2) possibly exacerbated by the COVID-19 crisis. This situation does not spare France. Even if it is a high income country with a fairly reputable health care system (3), the situation of child and adolescent psychiatry (CAP) in France is problematic, to say the least (4).

In France, the CAP care system is based on a territorialized offer, organized into “sectors” of around 200 000 inhabitants, comprising one or more medical-psychological centers (CMP for *Centre Médico Psychologique*), day hospitals and inpatient beds. The missions assigned to these sectors were set out in a circular dated March 16, 1972. These missions are very broad: prevention, diagnosis and treatment of mental disorders, coordination between the various actors involved in care. The particularly significant increase in demand for mental health care noticed since the end of the twentieth century (5), combined with the undersizing of sectors from the outset, have led in a gradual, then generalized overloading of the CAP care system by demand. As a result, waiting lists grew longer, sometimes exceeding an average of 6 months for an initial consultation appointment (4).

One-off attempts are regularly proposed at the national or regional level to overcome this unacceptable situation. None of them has changed the situation which continues to deteriorate.

The SFPEADA (French Society for Child and Adolescent Psychiatry and Allied Professions), founded in 1937, is one of the oldest national organization affiliated to th e IACAPAP (International Association for Child and Adolescent Psychiatry and Allied Professions). According to its statutes, SFPEADA is dedicated to “promoting and coordinating studies, research and training activities on mental, emotional and intellectual disorders in children and adolescents, on their treatment, as well as their prevention. It aims to bring together members of the various professions working towards this goal”. In this context, the SFPEADA decided in June 2023 to undertake an action research project aimed at specifying the contours of the French CAP in the second quarter of the 21^st^ century. More specifically, the aim of this project was to lay the foundations for a reflection on the place that CAP should occupy in France in the second quarter of the 21^st^ century, on its epistemology, its organization and the type of care it should be able to provide. As societal issues are at the forefront in such a context, the methodology adopted in this project was as global and comprehensive as possible, and combined bottom-up and top-down approaches.

As defined in (6), action research “seeks transformative change through the simultaneous process of taking action and doing research, which are linked together by critical reflection. Action research practitioners reflect on the consequences of their own questions, beliefs, assumptions, and practices with the goal of understanding, developing, and improving social practices. This action is simultaneously directed towards self-change and towards restructuring the organization or institution within which the practitioner works.”

In line with this definition, a 4-step action research project was designed: 1) videoconference interviews with “qualified persons” able to cast a critical eye (positive or negative) on the French CAP care system; 2) e-mailed open-ended questions to “intermediary bodies” involved in the issue of children’s and adolescents’ mental health, in line with a “deliberative democracy” perspective (“intermediary bodies” are typically associations and trade unions of professionals likely to interact with the field of child and adolescent mental health, scientific societies of the corresponding disciplines, parents’ associations, etc.); 3) production of a report based on the results of the two first steps and through a series of interactions between SFPEADA members; and 4) public presentation of the whole process during a one-day symposium at the French Ministry of Health.

## Methods

### Step 1: videoconference interviews with “qualified persons”

A purposive sampling maximizing heterogeneity and diversity led to 24 “qualified persons” being contacted. They had to be aware of the French CAP care system and could benefit from an experience of a foreign healthcare system, a mastery of an academic discipline (sociology, history, etc.), a personal experience as a patient’s parent, or the exercise of a profession or associative activity dealing with public health issues. Among them, 5 child and adolescents psychiatrists (CAPs) and 1 psychologist were currently working abroad (USA, Canada, Belgium, Spain, Switzerland, Iceland, 4 males, 2 females), 4 were social scientists (2 historians, 1 sociologist, 1 health economist, all females); 4 were parents of patients strongly involved in parents associations (2 females, 2 males); 7 were professionals close to the field (pediatrician, child welfare professional, school doctor, children’s judge, etc. 6 females, 1 male); 3 representatives of psychiatrists’ associations (2 males, 1 female), 1 representative of SFPEADA (male).

Twenty-three accepted to participate in a videoconference interview of about 30 to 60 minutes with fairly free instructions to present during around 20 minutes strengths, weaknesses, and necessary changes for French CAP care system. Then, all 3 authors (BF, AR and PE) who participated to most of the interviews, could ask questions to deepen or clarify certain points. All the interviews were recorded, transcribed with AssemblyAI (7), and then the corpus was coded with a thematic analysis following the 6 phases recommended by (8). BF and AR did the coding separately and confronted their outcome in a collaborative document. After that, all 3 authors met to determine the themes that were likely to organize best clusters of codes.

Natural language processing (NLP) was also used to analyze the corpus. The first NLP analyses were performed using R 4.3.2 software with “udpipe”, “tm” and “topicmodels” packages: after lemmatization, a text document matrix was estimated and analyzed with some hierarchical clustering and topic modelling applied using Latent Dirichlet Allocation. As these first NLP analyses were not conclusive, ChatGPT-4 was then used to triangulate the thematization done by the authors. Each interview was firstly coded by the IA, then all codes were gathered, and the IA was asked to thematize them. The prompts used (appendix 1) were inspired by the work of J Kriukow (9).

### Step 2: open-ended questions to “intermediary bodies”

Sixty-four “intermediary bodies” were contacted by e-mail. A cover letter (appendix 2) explained the objective and the context of the study, and the “intermediary bodies” were asked if they would agree to answer to two questions explicitly formulated: “1/ What should be the place and role of CAP in France in 2023 and in the years to come?” and “2/ What should be the place and role of a child and adolescent psychiatrist in this context?”. Thirty-four responded favorably, and 29 ultimately contributed to the study (7 trade unions or associations of psychiatrists, 2 trade unions or associations of psychologists, 2 trade unions or associations of speech therapists, 4 associations for mental health, 3 associations of patients’ parents, 1 associations for public health, 2 trade unions or associations of general practitioners, 1 association of hospital manager, 4 associations for child protection, 1 association for social welfare, 1 association of magistrate, 1 other) For reasons explained in the “results section”, one author (BF) coded the corpus and applied a thematic analysis. No triangulation with NLP was done.

### Step 3: synthesis

During a 3-day seminar, 8 members of the executive committee of the SFPEADA (6 CAPs, academics or not, working in big cities, smaller ones or rural areas, 4 males and 2 females), 1 psychologist (academic, male) and 1 speech therapist (academic, female) discussed each of the codes and themes that emerged from the previous two steps in order to provide “a reflection on the place that CAP should take in France in the second quarter of the 21^st^ century, on its epistemology, its organization and the type of care it should be able to deliver”. Thus, this reflection went from the succession of a bottom-up (step 1, 2 and 3) and a top-down (step 3) procedure.

A synthesis of these discussion was written by BF and successively sent for comments to the whole executive committee, the scientific committee and finally to all members of SFPEADA. After consideration of these comments, the synthesis has been marginally modified and sent to all the intermediary bodies participating in study 2 to collect their reactions that were added to the synthesis.

### Step 4: communication

In March 2024 the synthesis was sent to the media and to the ministry of health. The 23^rd^ of April, at the time of the “World Infant, Child and Adolescent Mental Health Day” (WICAMHD), a one-day symposium was organized in the ministry of health with a presentation of the synthesis, and several round-tables for debates between the different stakeholders of the field.

## Ethics

All participants, professionals and parents, gave verbal informed consent to participate and to the recording of the interviews. All transcripts were suppressed following acceptance of this paper.

According to Article R1121-1-II of the Public Health Code (Decree No. 2017-884 of May 9, 2017) this study does not require the approval of a personal protection committee (CPP).

## Results

### Step 1: videoconference interviews with “qualified persons”

The corpus of study 1 consisted in 126000 words. Three hundred and forty-two codes were consensually developed by BF and AR and condensed in 93 micro-themes. Saturation began after the analysis of 15 interviews and was complete after the 20th. Considering the structure of the interviews, the 11 themes that emerged from the micro-themes were nested in 3 domains: “Strengths”, “Weaknesses” and “Necessary changes” of CAP (figure 1).

**Figure 1.**
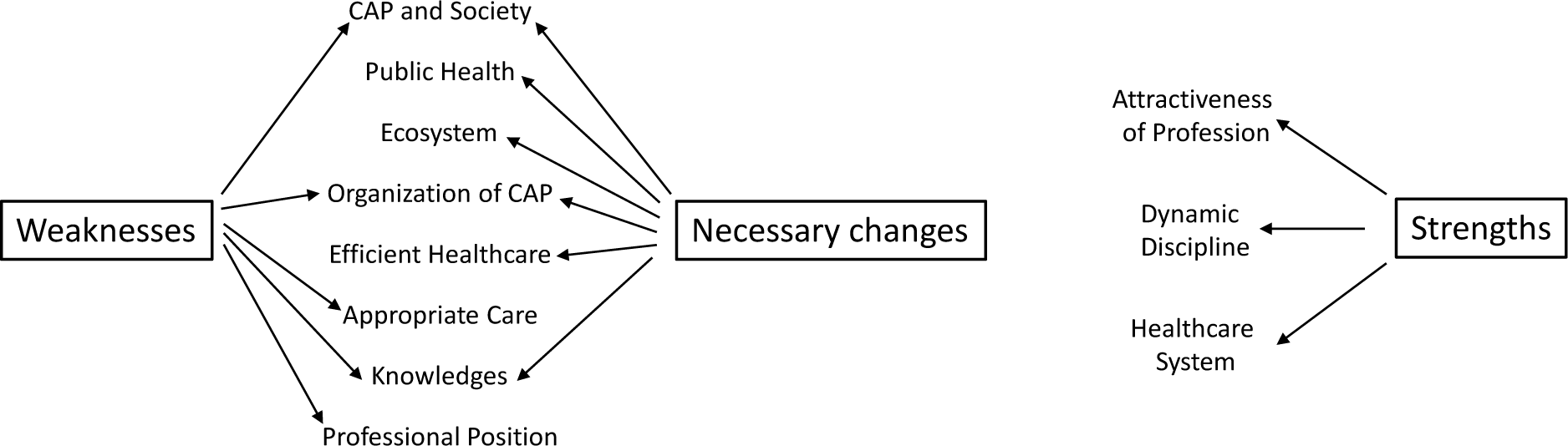
Thematic map synthesizing the analysis of study 1

**Domain “Strengths”** was marginally represented in the interviews, 3 themes were produced:

Dynamic discipline:training has improved; neurosciences changed the way we think about patients; early treatments are promising.

“We have a wide range of approaches, theoretical approaches, there’s the contribution of neuroscience, the contributions of neuroscience of course. From the pharmaco point of view too, it’s all very well, all the enrichment in any case. We have a wide range of frameworks, systems, techniques, integrative and transdisciplinary practices.” (S1, CAP) “we have nevertheless evolved and made progress in terms of prevention, early detection and treatment, which can be provided at an earlier stage.” (S1, CAP)

Healthcare system: organization into sectors reduces health inequalities; there is a comprehensive way to consider care.

“When it’s working well, I think that there’s an interesting territorial network of outpatient care between the CMPs and CMPPs, that it’s free, generalist, direct access, and that, for a form of health democracy, I think it’s something important.” (S2, CAP)

Attractiveness of profession: CAP is an intellectual profession which allows freedom of thought and practice; there is a diversity of models and practices.

“ I think it’s great, it’s a hyper-intellectual specialty.» (S3, paediatrician)

“And one of France’s strengths is the freedom to practice psychotherapy.” (S4, CAP, US)

“And I think that’s very positive. So there’s a great diversity of care on offer in France.” (S5, CAP, Iceland)

**Among domain “Weaknesses”**, 5 themes were produced:

<u>CAP and society</u>: there is an increase of demand and a massive shortage of offer; health inequalities are important; professionals are losing their legitimacy; inclusion, in particular at school, is a failure.

“Those who are able to finance a specialized public school increase their child’s chances of inclusion.

A few dominant figures in the voluntary sector, opinion leaders with specific socio-cultural and intellectual capital, steer public policy, illustrating a form of lobbying that does not benefit the most disadvantaged families.” (S6, Sociologist)

“My estimate is that in France, there are at least 200,000 children without a solution at home.” (S7, Parent)

<u>Knowledge</u>: there is a lack of research, in particular a lack of epidemiological data; the level of evidence of many treatments is poor; the over presence of psychoanalysis borders to obscurantism; the over presence of neuroscience borders to scientism.

“What I deplore today in these professions, because you’re talking about child psychiatry, is that the curricula for educators, social workers, psychologists, etc. are still full of psychoanalysis, and that it’s useless.“ (S8, parent)

“The social aspects are dismissed by Fondamental researchers and the new psychologists, who reduce most childhood disorders to a biological ethiology.” (S6, sociologist)

Organization of CAP: the system is too complex and rigid; there is no quality control; professionals are working in silos; many institutions work without any CAPs, in particular pediatric emergency department; professionals are in distress.

“Personally, when we were working on sectorisation and it was explained to me that there had to be three months’ residence in the right place before changing sector, etc., I have to admit that I fell off my chair a little, especially when it comes to children whose situation is very, very fluid.” (S9, administrator)

“So from the outside, for people who come into the system because they are ill, because they need treatment, understanding the system is extremely complicated. It remains opaque.” (S10, health economist)

Appropriate healthcare: waiting list are too long; CAPs are losing their time in non-medical activities; treatments are generally not appropriate; parents are in distress and made to feel guilty.

“Yes, I wanted to say that from the point of view of the reality principle, it’s no longer working. There are two responses to this: either we change clinical practice, or we say, indeed, there aren’t enough resources left for clinical practice as we want to continue to deploy it. Exactly. But once we’ve said that, what do we do about the other children on the waiting list? Do we put them all on Prozac or what?” (S11, school doctor instructor).

“If we go back to France and its policies, we can see that for several years now it has aimed to promote the increased role of family carers, particularly parents, which results in an increased burden on parents who have to rearrange their working hours or stop working and train to look after their children or spend a fortune.” (S6, sociologist)

Professional position: corporatism opposes too often psychologists to CAPs and compromises evolution, like sterile controversies that oppose ideologies and theories.

“We know that we’re going to have major difficulties on all sides, with the doctors’ unions, with the professional organisations of doctors, with the professional organisations of psychologists. We’ve already come under fire, saying ‘but we don’t want you to change the content of our duties at all’”. (S12, Parent)

“But what I see as the main stumbling block in France to the direct implementation of this is that we also have a certain number of teams that are completely sclerotic in their approach to care and ideology. In other words, when a new medical director takes over a unit, a CMP, or even an entire department, the team may be completely locked into archaic concepts that have no intention of being updated, and which are absolutely impossible to fire.” (S4, CAP, US)

**Among domain “necessary changes”**, 6 themes were produced: CAP and society: There is a need to take into account migrations and child abuse, which are important risk factors; the same is true with recovery and mental health literacy that are emerging concepts; CAPs should have a political role in prevention.

“I think that having precise guidelines from child psychiatrists can help a great deal in developing continuity in these pathways, in trying to put children in the same place. And then there are problems linked to very serious abuse or very complex migratory journeys that need to be dealt with in a fairly robust way. And there are also many children who refuse treatment in these services and who are not at all interested.” (S13, child protection services)

“It depends on social class, literacy and all that, but often, in fact, they don’t say what needs to be said because they don’t know what needs to be said at the right time, you see.” (S10, health economist)

Public health: need of an appropriate financial compensation of the different activities relevant to the field of CAP; of the development of a mental health information system; prevention is crucial in CAP; a strong political will is needed to achieve these goals.

“Oh yes, that’s perhaps an important factor. This computerization and automation […] we were left free to look after the patients and they were the ones who did virtually all the coordination.” (S14, psychologist, Spain)

“Outpatient child psychiatric consultations have been upgraded, so there are a lot of child psychiatrists working in the private sector too, so there are fairly easy links to the private, or I would say liberal, system. These nomenclature codes also made it possible to finance consultations with psychologists, to finance consultations with judges, with youth welfare services, so there was also a fairly extensive network of external therapists.” (S15, CAP, Belgium)

Ecosystem: multiple institutional partnership (school, special education, health authorities, judges, child protection services) is necessary; care itself must be considered on a systemic basis.

“What is the judge going to do with this expert report? And I would say that it’s in the regular interactions we can have, in the exchanges we can have, that we can reassure each other about both the scope of the mission, and what is expected, but I think that experts and judges need to talk to each other. It can’t just be a matter of writing up the expert opinion.” (S16, child judge)

“You see what I mean by that, prevention also means… because if we, as parents, don’t have the right support, and it’s also the fact that we know we can count on the child psychiatrist, at some point we also need to know that we’re not on our own. And that also helps because it keeps you going. You have to hold it together.” (S7, parent)

Knowledge: CAP requires multiples sources of theories and disciplines; research has to be promoted like the training of all the professional involved in the domain.

“I want there to be people from health, people from social services, the educational world and the school world, I want to mix social and health, by making them change roles, we have training techniques, it is very much based, it is a mixture of theory and experience so that people question themselves, in any case where we are talking precisely about posture, about authority, so that they realize what is happening between them, so that basically, they try to promote a change in the network.” (S17, CAP, Switzerland)

Efficient Healthcare: parents must be involved in care and considered as partners; treatments must be structured and transparent to patients and their family; CAPs have mainly to be managers, experts in diagnosis and treatment and have only a limited practice of psychotherapy; e-Health (online therapies, telepsychiatry) have a great potential.

“So first of all, on the quality of care, the care system, we have a problem with the lack of a culture of assessment, and here I’m going back to what xxxxx said, so double assessment of both symptoms, which means standardized data collection where the identification or diagnosis is formalized. This is very difficult to put in place in France, there’s still a lot of resistance, whereas in the United States or Canada, it’s part of the basis of any consultation.” (S4, CAP, US) “This led us to believe that it was important for child psychiatry to be able to focus on psychopathological issues and play a role both as an expert in diagnosis and as a coordinator of care, working with a multidisciplinary team” (S16, Child judge)

<u>Organization of CAP</u>: the role of each profession has to be clarified (for instance specificity of a CAPs versus a psychologists); there is a need of a graduation of care, level one must be de-medicalized; density of treatment must be considered in line with the severity of disease; out-patient treatment must be the rule; “In my opinion, there is a lot of room for improvement in the CMPPs, the CMPs, and the CMPs absolutely have to be structured so that they reach… because today, in France, for ADHD, there is no level 1, there is no level 2, there is no level 1. In practice, there is neither. So I’m calling for the creation of level 3s, so that level 3 forms level 2 and level 1.” (S8, parent) “But at some point, we’re going to have to coordinate and prioritize, and we’ll reassess the situation at a later date in the light of the choices we’ve made, and I think that’s important.” (S18, CAP) Interestingly, chatGPT-4 proposed a thematization close to the present one, 303 codes led to 11 themes (figure 2, see supplementary materials, appendix 3 for details).

**Figure 2.**
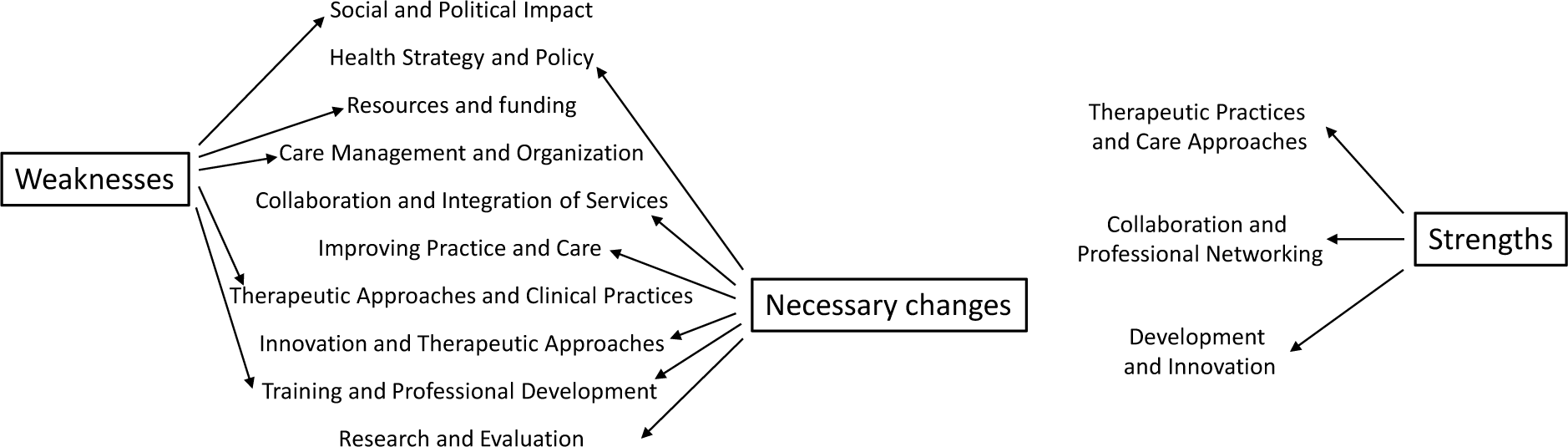
Thematic map synthesizing the analysis of study 1 (ChatGPT-4 analysis)

NLP was not conclusive: clusters proposed by hierarchical clustering (see supplementary materials, appendix 4 for an illustration) or by Latent Dirichlet Allocation were not interpretable and/or poorly relevant.

### Step 2: open-ended questions to “intermediary bodies”

Among the 64 intermediary bodies that were contacted, the 31 which contributed provided a very heterogenous content in quantity and quality, in particular concerning the relevance to the questions asked. The analysis of the 12600 words collected has thus to be considered cautiously and was done by one author (BF).

For question 1, 64 codes were developed, they were condensed in 23 micro-themes which end-up to 5 themes. For question 2, 43 codes were developed, condensed in 10 micro-themes which end-up to 4 themes. Results are presented in table 1.

**Table 1.** Thematic analysis of the open-ended questions to “intermediary bodies”. Question 1 (Q1): Role of child and adolescent psychiatry, Question 2 (Q2): role of child and adolescent psychiatrists.

|  |  |  |
| --- | --- | --- |
| Q1 | Around CAP | CAP, and prevention in particular, involve the society as a whole; institutional partnership must be the rule (school, judges, child protection services, etc.) |
| Q1 | Societal changes | Migrations, screen use, family involvement requires CAP to be highly adaptable. |
| Q1 | Knowledges | CAP is at the intersection of multiple disciplines. Training is of a particular importance for all professionals |
| Q1 | Healthcare provision | CAP is a medical specialty where there are diagnoses and treatments, pharmacological or not. CAP is multidisciplinary inherently, families are a part of the care. |
| Q1 | Healthcare system | It must be graduated, CAP is at level 2 and after; sectorization must be preserved; crisis unit and emergency department must be available. |
| Q2 | A coordinator | CAPs organize specialized care, coordinating a multidisciplinary team in conjunction with other professionals. However, they are not the only legitimate person for such a position. |
| Q2 | An expert | CAPs keep abreast of the latest research; they do in-depth interviews leading to reliable diagnoses and prescriptions; as consultant they give clinical advice to other professionals; they know psychotherapies. |
| Q2 | Multiple interlocutors | They interact with families and speak for the discipline externally. |
| Q2 | Responsible | For the quality and appropriateness of care, for ethical values and independence from the administration and fashionable theories. |

### Step 3: synthesis

From the results of study 1 and 2 and their intensive discussion during step 3, 5 themes have been proposed to summarize the whole process: “Society”, “Knowledges”, ”Care”, “Caregivers”, ”Care system” (figure 3.)

**Figure 3.**
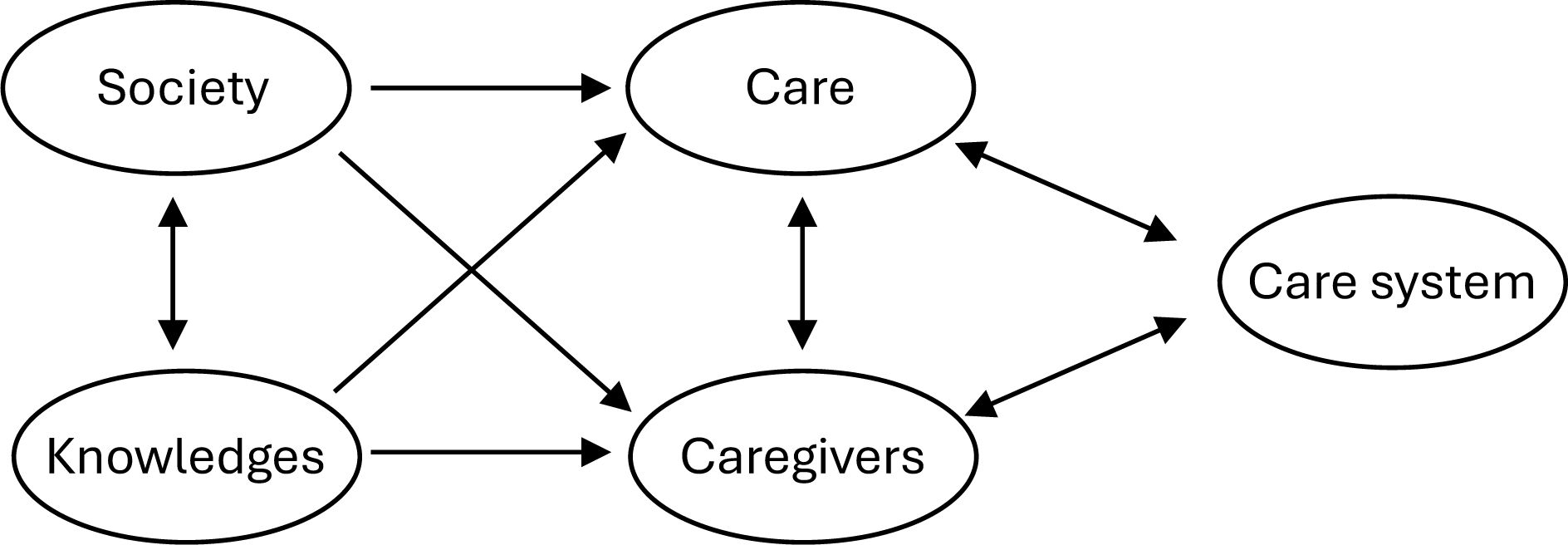
Thematic map synthesizing results from step 3.

A translation of the text that develops these themes to provide “a reflection on the place that CAP should take in France in the second quarter of the 21^st^ century, on its epistemology, its organization and the nature of the care it should be able to deliver” is proposed in the supplementary materials (APPENDIX .5). To make it easier to read and maximize its impact, this text is deliberately concise (14 pages, 5500 words including 32 references) and written in a non-technical language Step 4: communication

The text obtained in step 3 has been sent to a list of journalists specialized in medicine at the beginning of April 2024 to amplify the impact of the feedback conference which took place the 23^rd^ of April 2024 at the ministry of health. This conference was the occasion for a democratic debate between all stakeholders involved in child and adolescent psychiatry mental health and psychiatry.

Following the conference, the text has been sent to political parties, public health institutions and administrations. Regional health agencies, which are responsible for organizing the healthcare system at regional level, were key targets.

## Discussion

CAP cannot be considered independently from the society in which patients and their families live. Because of that, patients, parents, teachers, politicians, public health professionals and many others are legitimate to think about the role and the organization of CAP. Of course, professionals of the domain are legitimate too. Their experience is unique and particularly valuable when considering the complexity of situations they face every day. But, as professionals, they can be biased. Biased because of their training, their habits, their a priori concerning patients and their families.

In 2023, at a critical time for CAP, SFPEADA decided to highlight the position of CAP professionals on what should be CAP in the second quarter of the 21^st^ century. To minimize the potential biases that these professionals could have towards their own activity, an original methodology was adopted.

First, a bottom-up approach made it possible to collect the opinions and ideas of a wide range of stakeholders. In contrast, the second time of the research was more of a top-down nature.

Challenged by the stakeholders’ proposals, a group of CAP professionals, mostly child and adolescent psychiatrists, used their experience to produce a text gathering proposals that sound clinically relevant, feasible and sustainable in terms of health care organization and in line with patients, families and, more generally, societal expectations. Finally, again in a bottom-up perspective, the text was sent to the intermediary bodies involved in study 2 to collect their comments and feedbacks.

Among the strength of this work, there is first its “action research” positioning. The research part is designed to fit in optimally with the dissemination part. The overlapping of both has generated an undeniable burst of enthusiasm in the community. It was said that research was underway and that the results would be made public very soon. Attention was stimulated and light was projected on the event. Another strength concerns the method of analysis of study 1. A double triangulation was used: a classical one between 2 then 3 human beings and a second one between the 2-3 human beings and an AI. The coding and the thematization of chatGPT-4 were impressive. We noticed however some differences with the work of a human being. ChatGPT-4 was more neutral, closer to the corpus. BF and AR coding implicitly insisted on the more sensitive aspect of the corpus, as they had attended the interviews and remembered the interviewees’ emphases and intonations, so that their analyses appeared more forceful.

Among the weaknesses and limitation of the work, obviously study 2 is frail. Too few intermediary bodies sent a production that could be exploited. However, the content analyzed was rather homogenous and the coding quite straightforward so that results of study 2 do have an interest.

Another limitation comes from the composition of the group of professionals involved in step 3. Most were child and adolescent psychiatrists and many of them had an academic situation which is an obvious source of bias. However, the synthesis coming from step 3 has been extensively challenged, commented, and edited by an important number of SFPEADA’s members representing a vast number of professions and types of practice.

In the last years many important contributions have appeared which promote a new way to deal with mental health in a global perspective (1,10,11). What these papers, reports or action plans have in common is their focus on mental health much more than on psychiatry. Indeed, because prevention is the most efficient way to deal with CAP problems, promotion of mental health appears legitimately as a priority, and this even if the concept of mental health itself is not so clear (12). Some authors go very far in this direction, proposing for instance that “care is not contingent on a categorical diagnosis but aligned with the staging model of mental illness” (1). If categorical diagnoses are frequently criticized in CAP, in particular by some authors of this paper (13), we have been surprised by the importance that many parents of patients, teachers, judges, social workers and other professionals attach to psychiatric diagnoses. More generally, if there is no doubt that promoting mental health is a crucial issue for the next years and decades, it is just as important to set up an efficient psychiatric healthcare system. This is precisely what SFPEADA’s members had in mind with the present work: rethinking child and adolescent psychiatric care in collaborative way, in a period where most societies are particularly worried by mental health issues.

## Data Availability

Data produced in the present study are not available.

## Acknowledgements

The authors which to thank all parents, professionals and societies that participated to the 2 studies.

This paper has been supported by QUALab, the Qualitative & Mixed Methods Lab, a collaboration between the the Yale Child Study Center (New Haven, CT), and CESP, the Centre de recherche en Epidémiologie et Santé des Populations (Paris, France).

