## Appendix for "What child and adolescent psychiatry in France for the second quarter of the 21^st^ century? An AI-assisted qualitative action research study"

**Supplementary material**

**Appendix 1: ChatGPT-4 prompts (translated in English) that were used for the AI thematic analysis of study 1.**

***Prompt for coding:***

*Prompt 1*

You're a researcher. I'm now going to download an interview transcript and you're going to do what's called qualitative coding - in particular, initial coding, also known as open coding. The text is an interview transcript concerning French child and adolescent psychiatry. I want the codes to relate to the strengths, weaknesses and desired changes in French child adolescent psychiatry. The codes need to be detailed and descriptive. I want you to apply codes to sentences or parts of sentences and, later, when you draw up a list of codes, I want you to be able to tell me to which sentences or parts of sentences these codes have been applied. In other words, when I ask you to provide me with examples of quotations for the codes you create, I'd like you to be able to do so. The results should take the following form: the code, preceded by the word "code" and the code number with the code label (i.e. a synthetic description of what it represents) and specifying on the next line whether it relates to "strengths", "weaknesses", "desired changes" in French child adolescent psychiatry or, if it doesn't fit any of these categories to "another theme", then, on the next line, a slightly more detailed description of this code, preceded by the word "description", then on the next line one or more citation examples. Here's the text to code:

*Prompt 2 (if the number of codes is not sufficient, which is frequent):*

I would like more codes

***Prompt for thematization:***

*Prompt 1*

You are a researcher. Here is a list of codes that have been created during the qualitative coding of the content of several interviews relating to the changes needed to evolve French child psychiatry. Each code is presented on a line starting with the word "code" followed by the code number, then the code label, then the word "description" followed by a description of the code. Can you group these codes into several categories and sub-categories? these categories should be roughly comparable in size. Here are the codes:

*Prompt 2 and 3 have been used in the same way for strengths and weaknesses*

**Appendix 2. Translation of the covering letter sent to the "intermediary bodies" (study 2).**

Depending on the country and the period, child and adolescent psychiatry and child and adolescent psychiatrists have played very different roles.

In terms of the discipline itself, although it is universally recognized as a medical specialty, it has been able to take an interest in contexts as diverse as the psychiatric asylum, schools and special education, psychotherapeutic practice, pharmacology and neurobiology, pediatric emergencies, the treatment of psychological distress, social care for children with disabilities, and even the full realization of the potential of the younger generations.

While it is also widely recognized that the practice of child and adolescent psychiatry can only be conceived in the context of a team consisting of multiple and complementary areas of expertise, the role of the child and adolescent psychiatrist in this collective varies greatly from country to country. In emerging countries, the scarcity of child and adolescent psychiatrists means that they have to play a political, local, regional or national role in organizing care and preventing psychiatric problems in young people. In contrast, in some countries the child and adolescent psychiatrist is essentially an expert whose role is to make one or more diagnoses according to an official nomenclature and to propose treatment based on scientific data. This care is coordinated and provided by non-medical professionals. The situations in between are certainly the most numerous.

With these two observations in mind, we would like to hear your views on these issues. Responses can be short (a few lines) or long (no limit), the objective is that we can get as many points of view as possible. These will of course be anonymized during the analysis and when the results will be reported.

Question 1: What should be the place and role of child and adolescent psychiatry in France in 2023 and in the years to come?

Question 2: What should be the place and role of a child and adolescent psychiatrist in this context?

**APPENDIX 3. Thematization proposed by ChatGPT-4 (study 1)**

**Domaine: “Necessary changes”**

Theme 1. Health Strategy and Policy

*National and Strategic Development*: This sub-category includes codes relating to the need to develop a national strategy for child and adolescent mental health. It encompasses the development of comprehensive policies, the impact of legislation, adaptation to societal expectations, and the need for significant investment.

*Laws and Regulations*: This sub-category brings together codes that emphasize the importance of legislative and regulatory influence on child psychiatric practice. It concerns the balance between different psychiatric approaches and the integration of the rights of children and adolescents.

Theme 2. Training and Professional Development

*Training and Education of Professionals*: This sub-category includes codes that address the importance of continuing education and training of professionals in child psychiatry. It emphasizes the need for professional development, diversification of training and awareness-raising among professionals.

*Development of Specialized Skills*: Includes codes that emphasize the need to develop specialized skills, particularly in the diagnosis and management of complex and neurodevelopmental disorders.

Theme 3. Innovation and Therapeutic Approaches

*Diversification of Therapeutic Methods*: This sub-category brings together codes that call for greater diversity in therapeutic approaches, including the integration of psychoanalysis, neuroscience and non-medicinal methods.

*Use of Technology*: This sub-category brings together codes that highlight the growing importance of, and the need to properly supervise, the use of digital tools and artificial intelligence in diagnosis and case management.

Theme 4. Collaboration and Integration of Services

*Intersectoral Coordination*: Includes codes highlighting the need for better coordination between the different sectors involved in child psychiatry, as well as the integration of mental health services for more effective care.

*Collaboration with Families and the Community*: This sub-category includes codes that emphasise the importance of partnership with families and the need for community-based approaches to treatment.

Theme 5. Improving Practice and Care

*Improving Accessibility and Continuity of Care*: This sub-category includes codes that address the need to improve accessibility and continuity of care in child psychiatry, including the opening of new beds and flexible care schedules.

*Holistic and Personalised Approaches*: This sub-category includes codes that emphasise the importance of adopting holistic and personalised approaches to effectively meet the individual needs of patients.

Theme 6. Research and Evaluation

*Need for In-Depth Research*: This sub-category brings together codes that emphasise the need for in-depth research to understand the specific needs of protected children and to evaluate current practice.

*Evaluation and Improvement of Practice*: Includes codes that emphasise the need for continuous evaluation and improvement of child psychiatry practice, based on evidence and rigorous research.

**Domain “Weaknesses”**

<u>Theme 1. Resources and funding</u>

*Lack of Resources and Funding*: This sub-category includes codes that address the general lack of human, material and financial resources in child psychiatry. It addresses issues such as under-funding, lack of child psychiatrists, and lack of appropriate structures.

*Infrastructures and Equipment*: This sub-category groups together codes relating to specific challenges concerning physical infrastructures, such as the lack of hospital beds, security problems in establishments, and structural difficulties in emergency services.

<u>Theme 2. Training and Professional Development</u>

Lack of Training and Expertise: This sub-category covers codes which highlight the lack of specialised training and professional skills, particularly in the diagnosis and management of complex cases. It also includes problems of accessibility to training and the need to improve education in child psychiatry.

Recruitment and Professional Mobility: Here are grouped the codes that address the difficulties related to recruitment, retention and mobility of professionals in child psychiatry. This includes medical demography, the attractiveness of the sector and future recruitment challenges.

<u>Theme 3. Therapeutic Approaches and Clinical Practices</u>

*Clinical Challenges and Case Complexity*: This sub-category groups together codes that deal with the increasing complexity of clinical cases, the prevalence of certain pathologies and the challenges posed by marginal cases. It also includes critiques of current therapeutic models.

*Criticisms of Current Therapeutic Approaches*: This sub-category includes codes which criticise current clinical practices, highlighting the need for innovation and diversification of therapeutic approaches, and pointing out the limitations of existing models.

<u>Theme 4. Care Management and Organisation</u>

*Lack of Co-ordination and Continuity of Care*: This sub-category groups together codes that address fragmentation and lack of co-ordination in the care pathway, difficulties in transition between different phases of life and challenges in managing care.

*Management and Supervision Challenges*: This sub-category includes codes relating to management and supervision issues within child psychiatry settings, including resistance to change, public policy limitations and supervision and evaluation challenges.

Theme 5. Social and Political Impact

*Socio-economic and Political Issues*: This sub-category includes codes that address socio-economic and political issues affecting child psychiatry, such as inequalities in access to care, the impact of prejudice and misconceptions, and the lack of political will for meaningful reform.

*Stigma and Awareness*: This sub-category groups together codes that address the stigma of mental illness and the lack of awareness of these issues, highlighting the need for better communication and understanding between health professionals and families.

**Domain "Strengths"**

Theme 1. Therapeutic Practices and Care Approaches

*Diversity and Evolution of Therapeutic Approaches*: This sub-category includes codes that highlight the diversity and progress of therapeutic approaches in child psychiatry. It highlights the adoption of innovative practices, the broadening of diagnostic criteria, and the integration of various methodologies, including contributions from the neurosciences.

*Patient and Family Centred Approaches*: This sub-category groups together codes that emphasise the importance of a personalised, patient-centred approach, taking into account the impact and role of the family in the care process. It also emphasises the need to adapt treatments to the specific needs of each patient.

Theme 2. Collaboration and Professional Networking

*Interprofessional and Multidisciplinary Collaboration*: This sub-category includes codes that value collaborative and networked working between different healthcare professionals. It highlights the importance of shared expertise, cooperation between the various players in the sector (child psychiatrists, psychologists, educators, etc.), and the multidisciplinary approach.

*Intersectoral and Community Collaboration*: This sub-category groups together codes that emphasise the importance of collaboration between different sectors (education, health, social services) and with the community. It highlights the benefits of integrating child psychiatry services into the social and educational fabric, and of the community approach.

Theme 3. Development and Innovation

*Progress and Innovation in Child Psychiatry*: This sub-category includes codes that highlight advances and innovations in the field of child psychiatry. It covers the progress made in destigmatising mental health, the importance of prevention, and the positive contributions made by parents' and patients' associations.

*Valuing Experience and Expertise*: This sub-category groups together codes that highlight the expertise and dedication of child psychiatry healthcare professionals. It recognises their experience, their ability to adapt to complex situations, and their crucial role in the development of the field.

APPENDIX 4. Hierarchical cluster analysis of the text*document matrix of study 1 corpus (Ward distance, documents consist in 20 consecutive tokens).

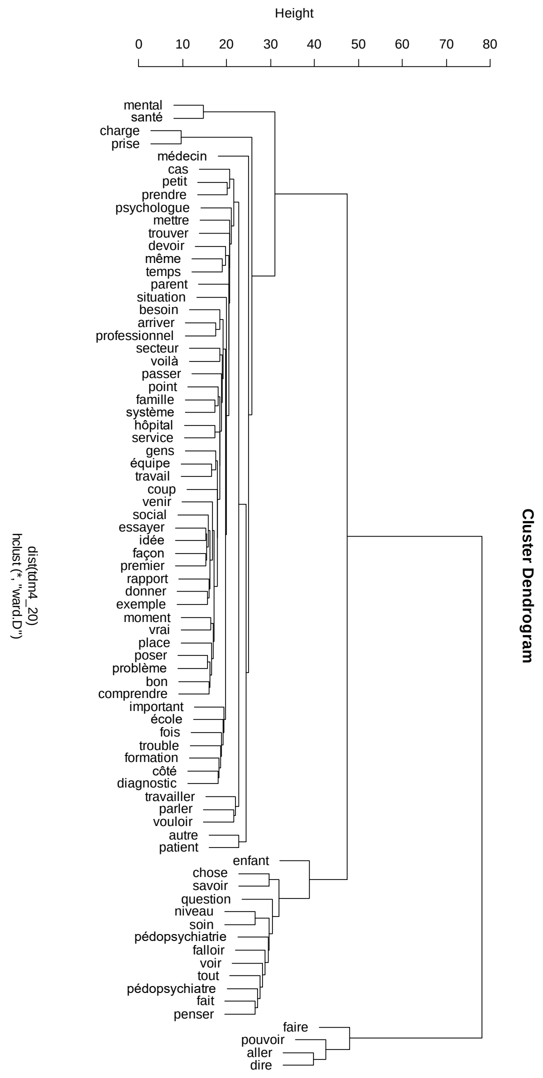

APPENDIX 5. Translation of the text “What child and adolescent psychiatry in France for the second quarter of the 21st century?” sent to the media, political parties and public health institutions.

Here's the translation of the entire document from French to English:

**Introduction**

The French Society of Child and Adolescent Psychiatry and Associated Disciplines (SFPEADA) organizes on April 23, 2024 (World Mental Health Day for Children and Adolescents) the "Conference on the Reestablishment of Child Psychiatry." The purpose of this conference is to reflect on the role child psychiatry should play in 21st-century France, its organization, and the nature of the care it should be able to provide. At a time when much is said about mental health and little about psychiatry, when the demand for psychiatric care for children and adolescents is increasing while the number of child psychiatrists is decreasing, and when new clinical concepts of very different nature (e.g., "neurodevelopmental disorders" versus "recovery") are emerging, SFPEADA found it appropriate to take the time to reflect to propose a rebuilding of the discipline.

To carry out this reflection, SFPEADA has commissioned two studies:

The first study is scientific. It was based on interviews with "qualified personalities" who can provide a critical (positive or negative) perspective on the French pediatric psychiatric care system. In addition to the knowledge of this system, this qualification could come from experience in a foreign care system, mastery of an academic discipline (sociology, history, etc.), personal experience as a parent of a patient, practicing a profession, or participating in an association open to public health questioning.

The second study was conducted among "intermediate bodies" ("social groups located at an intermediate level between the individual and the state, independent and autonomous, naturally formed or by deliberate agreement to achieve a collective objective") involved in the issue of mental health for children and adolescents. This study is more in line with a "deliberative democracy" perspective.

The analysis methodology and the results of these two studies are proposed in a longer version of this report. To summarize, a thematic analysis was performed which led to 113 themes grouped into a limited number of "major themes," five of which can be retained:

- Society
- Knowledge
- Care
- Caregivers
- Organization of the healthcare system

During a 3-day seminar, 8 members of SFPEADA (hospital-university or hospital child psychiatrists practicing in a metropolitan area or rural environment, as well as two professionals from associated disciplines) freely discussed each of the 113 sub-themes resulting from studies 1 and 2 to derive a professional view on the desirable evolution of this discipline over the next 25 years.

A synthesis of these discussions was then written. It thus proceeds from a double movement: "bottom-up" in collecting points of tension and necessary changes, "top-down" in formulating the analysis and proposals.

This synthesis was subsequently proposed for discussion to the board of directors and the scientific council of SFPEADA, then to all members of the society. This report takes into account these discussions. Parallelly, an article presenting all these works was written for submission to an international peer-reviewed journal. Furthermore, this report will be sent to all the intermediate bodies that participated in study 2 so they can discuss its content.

**Discussion of Theme 1: "Society"**

Children and adolescents evolve in an environment that has changed significantly in recent decades. Some of these changes are positive: reduced stigmatization of certain pathologies, particularly autism (1); awareness of the importance of combating domestic violence, an undisputed risk factor for numerous psychiatric pathologies (2). However, other changes may have a negative impact: increased academic pressure (3); concerns about the future, particularly in relation to the climate crisis (4); identity questions (5); risks associated with new communication technologies (6). The COVID epidemic has also significantly affected the mental health of adolescents (7). These changes might explain the observed changes in the clinical expression of disorders presented by young people in recent years, noted by a large number of clinicians.

Regarding families, an increased awareness of the need to address psychiatric problems in youth probably explains the noticeable increase in the demand for care observed since the late 20th century (8). Thanks to the internet, some families are now informed of the main scientific knowledge about their child's pathology with the desire to be fully involved in care, which may alter the relationship between caregivers and families. There is also a greater demand for quick and effective responses to any care request.

On a broader societal level, the boundaries defining what may constitute a "disease," a "disability," or a mere "identity" are shifting significantly without the various stakeholders being fully aware and without clear and consensus-based definitions being provided by each group to these different concepts. Therefore, the very goal of child psychiatric care deserves to be better clarified: sometimes reducing symptoms, but perhaps more importantly, helping the young person regain as full a place as possible at school and in society (9).

Still on a societal level, although the stigma associated with certain diseases is regressing, the issue remains central and affects not only the patients and their families but also the caregivers themselves (10).

**Discussion of Theme 2: "Knowledge"**

Because child psychiatry deals with a developing being indissolubly linked to the family structure in which they live and the society that surrounds them, this discipline necessarily calls upon multiple sources of knowledge.

Some of these knowledges are described as fundamental. For instance, those derived from neuroscience, cognitive sciences, and developmental psychology are all essential for caregivers involved in pediatric psychiatric care, given that a newborn, a child, and an adolescent have brains that undergo considerable transformation. An understanding of the insights from neuroscience is especially necessary for child psychiatrists because they are physicians and thus custodians of knowledge related to the functioning of the human body, and they need to understand the mechanisms of action of the medications they prescribe. Although neuroscience has made spectacular progress in recent decades and helps to better understand some pathological processes, it is observed that it still has only limited practical applications (11,12), except in the discovery of rare organic pathologies with psychiatric expression that are accessible to specific treatment.

For a long time, psychoanalysis was a nearly exclusive source of fundamental knowledge in child psychiatry. It is now particularly criticized. In some respects rightly so (some psychoanalysts have blamed parents in a caricatured, guilt-inducing, and erroneous manner for their children's problems) but also wrongly in some respects. Indeed, psychoanalysis can also help some caregivers to better understand complex and painful clinical situations they face. Moreover, psychoanalysis has enabled and continues to enable the development of care practices that have proven their effectiveness, such as in anxiety-depressive disorders in children and adolescents or borderline personality organizations

(13). Today, in many clinical situations, it is important not to oppose psychoanalytic and behavioral therapies but rather to think about how psychotherapeutic practices, which also include family therapies, dialectical emotion therapies, relaxation, etc., can be coordinated with each other and respond to the particular situations to be managed. Let us take this opportunity to remind that contrary to what is sometimes alleged, the training of interns in child psychiatry in 2024 in France only marginally calls upon the psychoanalytic corpus.

The social and human sciences in general can be summoned as useful knowledge for practicing child psychiatry. This includes sociology (for a better understanding of society's influence on the structuring of the family and the role of schools, for example), anthropology (to grasp the role of culture or even spirituality, particularly in a context of migration), history (to learn to put into perspective the dominant knowledges at a given moment), and economics (to think about efficient care).

Knowledge is not only fundamental; it is also statistical. Clinical research and epidemiology are essential for establishing rational strategies for prevention and care; they both contribute to the emergence of an evidence-based medicine (EBM). However, it should be recalled that EBM suggests that medical decision-making should be the result of statistical study results, patient preferences, and the clinical experience of the doctor who will contextualize the statistical results of scientific studies to the unique clinical situation they encounter (14). This tripartite determination of medical decision-making is particularly important in child psychiatry because of the wide variety of clinical situations encountered.

Child psychiatric knowledge also relies on a semiotic and nosographic corpus (nosography is a methodical classification of diseases). This corpus is particularly complex due to the strongly evolving nature of behaviors and psychic functioning in developing humans. For example, depressive symptomatology does not manifest the same way in a child at ages 1, 7, or 16. Therefore, and because of a lack of convincing pathophysiology of pediatric psychiatric disorders, the nosography of these disorders is still very fragile; this is evident when considering the field of perinatology. American

(DSM-5) or international (ICD-11) classifications are often considered by non-specialists as "bibles" presenting psychiatric disorders with the greatest scientific precision as if they were in an almost definitive form. Nothing could be less certain. These classifications are certainly useful for facilitating research and the modalities of care reimbursement by insurance; however, they are evolving. To realize this, one only needs to look at the Research Domain Criteria (RDoC) project developed by the American National Institute of Mental Health (NIMH), which proposes a nosographic approach completely different from that of the DSM-5 (15).

Thus, there is a multiplicity of knowledges that constitute child psychiatry. Caregivers must be fully aware of this. It is on this condition that they will be effective in their care, humble and clear about their limitations, flexible, and ready to adapt to different patients and teams, as well as to the evolution of knowledge and organizations. Refusing to see this is to take the risk of scientism or obscurantism, two equally dangerous pitfalls that daily threaten the caregiver looking for easy solutions.

To ensure these knowledges fulfill their role, it is necessary to promote research—all research, whether fundamental or clinical, quantitative or qualitative (16). This research needs to be carried out as close as possible to the point of care, and in collaboration with families. This will not only ensure their relevance to tomorrow's clinical practices, it will also facilitate their acceptance by caregivers.

Research is essential, but so is training. The sharing of knowledge between the various professions involved in child psychiatric care, but also with judges, teachers and all professionals involved in children in distress, is necessary to build up a common language and culture, essential foundations for better understanding and coordinating each other's decisions.

**Discussion of Theme 3: "Care"**

Child psychiatric care is overwhelmingly multidisciplinary in most situations. The main professions involved will be addressed in Theme 4; the institutional organization enabling this multidisciplinarity will be detailed in Theme 5.

These treatments must be efficient.

Efficiency first means that they must be effective. In medicine, adherence to Evidence-Based Medicine (EBM) principles is generally a guarantee of the efficacy of the treatments provided. In child psychiatry, however, this principle must be viewed with some nuance. Indeed, psychotherapeutic treatments play an essential role, and unlike drugs, their efficacy lends itself less readily to a traditional statistical approach like randomized controlled trials (17). Furthermore, conducting a scientifically valid study requires significant funding. For drugs, this funding is provided by pharmaceutical companies. For non-drug treatments, there is no equivalent, partly explaining why many treatments are not and cannot be validated in the strictest sense. This does not mean, however, that they are ineffective. Too strict an adherence to the EBM approach can lead to troubling public health situations, such as the excessive use of psychotropic drugs in treating children and adolescents. In all cases, as mentioned earlier, EBM inherently includes the patient's (and here, their family's) preferences and the clinical experience of the physician.

Efficiency also means that these treatments must be economically sustainable. The concept of treatment density is essential here. It is crucial to tailor the volume of treatment (e.g., frequency of sessions) to the pathology being treated and its severity. It is also essential to regularly assess the clinical progress of patients with the potential need to adjust or change the therapeutic approach.

There is an expectation from families that these treatments be transparent (i.e., families should know precisely what is being offered to their child) and structured (i.e., there should be clarity in how the different treatment modalities are coordinated and planned). These expectations are legitimate.

More broadly, the necessary involvement of families in child psychiatric care is evident both due to societal expectations and for clinical efficacy reasons. Approaches based on parental guidance, for example, are recognized for their high efficiency and would benefit from being more widely developed. The involvement of parent-peers in care is also an interesting avenue to explore.

The issue of family involvement is not straightforward, however. Domestic violence and abuse are among the primary risk factors for psychiatric disorders in children and adolescents (2). Hence, parents are sometimes, at least in part, the source of their child's disorders. It might be tempting to imagine that there are 'bad' parents who should be excluded from care and 'good' parents who are essential to the same care. The reality does not conform to this simplistic dichotomy. There is a continuum of good/poor treatment that lies at the heart of the complexity of clinical practice in child psychiatry. Ambivalence is inherent to all forms of parenting. It is thus challenging to define simply and systematically what the role of parents in child psychiatric care should be. This role must always exist—it is a given—but it must also be rethought according to the specificities of each family's dynamics.

In the second quarter of the 21st century, care cannot be conceived without considering e-health. There is a significant potential for progress if we learn to tame these new tools and use them appropriately. E-support-assisted therapies have already proven their interest in well-conducted studies (18). Teleconsultations can be useful both for patient follow-up and for promoting "indirect clinical practice." This latter consists of advising less medicalized structures during consultations or synthesis meetings conducted remotely.

E-health tools will also play a major role in the prevention of child psychiatric disorders (screening for disorders by families or by adolescents, improving mental health literacy, etc.). More generally, preventive actions are at least as important in child psychiatry as in the rest of medicine (19). Primary prevention (preventing pathologies from occurring) is primarily a matter of political decisions, and the role of child psychiatry in such a context is delicate. It can provide advice but should not be the main actor, both for reasons of efficacy and legitimacy, and even ethics (in the past, psychiatry has promoted social norms that have proven disastrous).

Discussion of Theme 4: "Caregivers"

If child psychiatric care is most often multidisciplinary, the various professions it calls upon are not always well identified by families and sometimes even by professionals themselves.

The child psychiatrist is a physician. He or she rarely performs clinical examinations but must nevertheless retain knowledge of somatic medicine, as this guarantees the safety of the drug prescriptions they make and underpins part of their legitimacy in the eyes of families and society. The child psychiatrist diagnoses psychiatric conditions, which is also what families and society expect from them. Child psychiatrists are sometimes reluctant about this diagnostic injunction. This reluctance is understandable when one recalls the fragility of psychiatric nosographies; it is even more understandable when one remembers that a child or adolescent is developing and that a diagnosis that seems obvious one day may be invalidated a few years later. This is the case for most disorders, including those that appear to be the most stable over time, such as autism spectrum disorders (20).

A child psychiatrist undergoes lengthy training—11 years post-secondary education. Hence, they are regarded as an expert in their field, and possibly for this reason—and perhaps also due to the aura of authority that surrounds the medical profession—they often coordinate the care provided to patients and their families. This is certainly the case in France and seems quite consensual, but this is not the case in all countries, and in reality, nothing obliges a child psychiatrist to occupy such a position. When this is the case, they should benefit from appropriate training, as nothing in their 11 years of education prepares them for this.

Child psychiatrists are often called upon to represent child psychiatric care to various institutions also dealing with children in distress or difficulty (national education, institutions dealing with disability or child protection, for example). This task is considered important, although in several countries, this activity is delegated to para-medical professions. Indeed, child psychiatrists need to preserve their time because the demographic of the profession does not at all match the needs of the population, and this is unlikely to improve significantly in the coming decades. The time for diagnosing, assessing, and reassessing care must be prioritized. A limited psychotherapeutic activity can also be preserved; it allows the child psychiatrist to keep in mind the complexity of clinical situations.

The initial training of child psychiatrists has recently changed; it is unquestionably of better quality and is now compatible with training programs in most European countries. However, continuing education is still largely improvable, even though it is of utmost importance given the significant changes the discipline is currently undergoing. It is also important because of a frequent tendency toward specialization of clinical practices over time, which harms the adaptability of the clinician. As with all medical specialties, the question of periodic re-certification is anticipated in regulations, desired by many caregivers, but not yet implemented.

Finally, the child psychiatrist is considered a guarantor of care—ensuring its relevance, quality, and ethics. For this, the child psychiatrist must maintain independence from actors likely to interfere with the care, whether they come from the administration or the judiciary, for example.

Psychologists have an essential place in child psychiatric care. They are likely to perform comprehensive assessments, including observations, clinical interviews, and the administration of psychological tests. They respond appropriately to the needs of the child and adolescent, relying on various therapeutic approaches. They participate in prevention and education activities with families and educational professionals to promote a better understanding of the psychological disorders of children and adolescents. The existence of specialization in the training curriculum of psychologists (clinical psychologist, neuropsychologist, developmental psychologist) is potentially problematic when it comes to caring for children or adolescents, as all three facets of psychology are highly useful in this field. This is all the more true as the content of training can vary significantly from one university to another. A more significant presence of psychologists in child psychiatry is certainly desirable, particularly due to a foreseeable de-medicalization of some care structures (see Theme 5). The creation of a child psychologist curriculum or a doctoral exercise program in psychology would facilitate such a transition, with the sensitive issue of the place of these professionals within an institutional hierarchy.

In terms of speech therapists, psychomotor therapists, and nurses, their role is also crucial. The university-level consolidation, or at least a reinforcement of the university nature of their training, is essential, with, as a corollary, the proactive development of research activities. The creation of a status of nurse specialized in psychiatry, following the model of nurse anesthetists (1 year of training after at least 5 years of practice), should be considered to give these caregivers, accustomed to the hospital setting, more responsibilities. This status would, of course, be different from that of advanced practice nurses (APN), a profession of great potential interest but which imposes a training difficult to implement (2 years, often far from home and family), and whose job profiles are still to be clarified.

Discussion of Theme 5: "Healthcare System"

The French child psychiatric care system is often considered complex, even incomprehensible (21). This is primarily the opinion of the families who use it, but also of many professionals who operate within it.

Beyond its structural complexity, the system is currently suffering from saturation, which prevents it from adequately responding to many perfectly legitimate care requests. Occasional adjustments are regularly proposed, generally within the framework of calls for projects, but the improvements observed are random and at best modest. Quite profound changes are therefore likely necessary; these will require considerable efforts both politically and administratively, as well as from the caregivers themselves.

It is customary to organize a healthcare system into levels of specialization. Regarding child psychiatric care:

- Level 1 would logically involve general medicine but also, and especially, schools, a critical environment for most children and adolescents. The question of training these various actors on issues raised by child psychiatry appears essential; it is currently considered largely insufficient.

- A Level 1bis should be considered. This would involve providing families and patients with a single, simple, and rapid non-medicalized access point structured by age groups, for example: “adolescent homes,” “family and children's homes,” and “first 1000 days homes” (to be necessarily thought of in conjunction with maternal and child health centers and early childhood action medico-social centers). Professionals from Level 2 would consult in these structures (possibly remotely) to help prioritize young people for whom further investigations might be necessary. Specifications would formalize the role of these structures and the modalities of interaction with adjacent levels, especially the educational environment.

- The core of Level 2 would correspond to the current sector of child psychiatry whose territorial network is an undeniable asset to reduce, as much as possible, the inequalities of access to care—a perennial problem of mental health care, particularly for children and adolescents. Associations such as the current medico-psycho-pedagogical centers would also be part of this Level 2; the same goes for private practices of child psychiatry, psychology, speech therapy, or psychomotor therapy. The potential referral of a patient from Level 1bis to care relevant to Level 2 would be based on the severity of the underlying pathology and the required density of care. The child psychiatry sector is intended to take care of the most severe cases. The question of specializing the care offered in the sector poses itself: by age groups, this certainly makes sense; by type of pathology (neurodevelopmental disorders, trauma, or eating disorders for example) or by type of environmental factor (migrant children) it is debatable. Indeed, such segmentation, while it rests on clinically relevant dimensions each requiring specific care, risks losing the necessary global vision of the patient. The problems of these patients can rarely be summarized by a single psychopathological dimension; the omnipresent notion of comorbidity is there to remind us of this.

- Level 3 would correspond to expert centers, university hospital services, and certain highly specialized units. This level of care should be exclusively reserved for the most complex situations. A frequently observed pitfall at this level is that highly specialized care proposals are made, although these are not generally available in reality. This sometimes creates frustration for both the parents and the downstream teams and can also be a source of inefficiency (22). It is therefore suggested that actors at Level 3 should not only play a simple role of expertise but also engage themselves in care.

- Crisis units and emergency reception are, of course, necessary. However, everything must be done to reduce their role as much as possible. Anticipated care is more efficient than care given in urgency or crisis situations; moreover, this type of care is particularly taxing for the teams, families, and patients. Currently, the expressed need for crisis and emergency reception units is linked to the inability of sectors, especially CMPs, to respond to the demand for care. Addressing this problem will, in fact, reduce the need for emergency and crisis reception, without eliminating it.

The French psychiatric sector has always been concerned with “going towards” patients and their families, conducting interviews in living environments, which decentralizes clinical viewpoints and may strengthen the therapeutic alliance and defuse difficult situations (23). In this context, if mobile teams are clearly to be encouraged, they can, however, only be conceived as a supplemental tool (except under specific local circumstances).

Organizing the healthcare system cannot be thought of other than locally. It is illusory to imagine that a single model could be applied across the entire French territory. The second quarter of the 21st century will still see territories better endowed (metropolitan areas close to university hospitals, for example) than others (particularly rural areas). In these latter, the principle of reality will force the maximum development of indirect clinical practice, mobile teams, and task delegation.

Organizing a healthcare system also means ensuring that everyone's time is used to the best advantage. Professionals whose  qualifications are rare and costly should not be used for tasks that do not involve care per se. This is typically the case with the recurring collection of indicators or administrative information, which has the primary consequence of lengthening waiting lists for welcoming patients and their families. The rationalization of institutional and cross-institutional meetings is also an efficiency issue. These meetings are desired and desirable because the link between professionals, teams, and institutions allows overcoming divisions too often observed in practice. The risk, then, is to prioritize this type of activity over care itself, sometimes more emotionally demanding. A balance must be found, remembering that care must remain at the heart of the caregiver's profession. This balance can be favored by a more appropriate pricing of clinical and non-clinical activities. Consultations, therapy sessions, institutional meetings, cross-institutional meetings, indirect clinical activity, home visits, hospital care, whether in a liberal or institutional setting: the remuneration of these acts must be thought out so that the whole is as efficient as possible, and none should be neglected.

Efficient care is not incompatible with caregivers who are fulfilled in their practice. Professional fulfillment is even a necessary condition for efficient care. This fulfillment is the condition for attracting people to these professions, which is currently lacking. Remuneration is certainly a key element, but it is not the only parameter. The freedom to think (not to suffer from more or less arbitrary ideological dogmas in the modalities of care to be provided), not to be considered as a simple interchangeable service provider, to benefit from a certain intellectual stimulation (regular training, moments of in-depth clinical reflection), and to have the means to provide at least acceptable care—this is what could, above all, give the necessary momentum to outline the contours of a child psychiatric care system.
